# Exploring patient experiences and concerns in the online Cochlear Implant community: a natural language processing approach

**DOI:** 10.1101/2022.03.22.22272761

**Authors:** Christopher Y.K. Williams, Rosia X. Li, Michael Y. Luo, Manohar Bance

## Abstract

**Importance:** There is a paucity of research examining patient experiences of cochlear implants, with existing studies limited by small sample sizes and closed question-answer style formats.

**Objective:** To use natural language processing methods to explore patient experiences and concerns in the online cochlear implant (CI) community when in conversation with each other.

**Design:** Cross-sectional study of the online Reddit r/CochlearImplants forum. No date restrictions were imposed; data were retrieved on 11 November 2021 and analysed between November 2021 and March 2022. The following details were extracted for each post: 1) original post content, 2) post comments, and 3) metadata for the original post and user.

**Setting:** Online Reddit forum r/CochlearImplants.

**Participants:** Consecutive sample of all users posting on the r/CochlearImplants forum from 1 March 2015 to 11 November 2021.

**Main Outcomes and Measures:** Natural language processing using the BERTopic automated topic modelling technique was employed to cluster posts into semantically similar topics. Topic categorisation was manually validated by two independent reviewers and Cohen’s Kappa calculated to determine inter-rater reliability between machine vs human and human vs human categorisation.

**Results:** We retrieved 987 posts from 588 unique Reddit users on the r/CochlearImplants forum. Posts were initially categorised by BERTopic into 16 different Topics, which were increased to 22 Topics following manual inspection. The most popular topics related to CI connectivity (n = 112), adults considering getting a CI (n=107), surgery-related posts (n = 89) and day-to-day living with a CI (n = 85). Topics with the most comments included Choosing cochlear implant brand (mean = 12.9 comments) and Adults considering getting a CI (mean = 12.2 comments). Cohen’s kappa among all posts was 0.62 (machine vs human) and 0.72 (human vs human), and among categorised posts was 0.85 (machine vs human) and 0.84 (human vs human).

**Conclusions and Relevance:** This cross-sectional study of social media discussions amongst the online cochlear implant community identified common attitudes, experiences and concerns of patients living with, or seeking, a cochlear implant. Our validation of natural language processing methods to categorise topics shows that automated analysis of similar Otolaryngology-related content is a viable and accurate alternative to manual qualitative approaches.

**Key points:** *Question:* What are the main patient experiences and concerns reported by the online cochlear implant (CI) community?

*Findings:* In this cross-sectional study of 987 posts from the Reddit forum r/CochlearImplants, we employed and validated a natural language processing approach to identify a variety of common themes discussed by current and prospective CI users. These ranged from CI connectivity and day-to-day living with a CI to queries about choosing a CI brand and post-op expectations.

*Meaning:* Analysis of social media using automated topic modelling techniques is a viable and insightful method to explore CI user experiences.

## Introduction

Over the past few decades, the availability of cochlear implant (CI) surgery has increased exponentially, with an estimated 736,900+ devices implanted worldwide.^1^ Several factors have contributed to this increase, including advancements in surgical techniques and device engineering, more widespread screening for hearing loss among children, and broadening of selection criteria for cochlear implantation.^2–4^ Despite this, the relative uptake of CIs among patients eligible for surgery remains poor, with rates of utilisation/provision as low as 6% in the United States.^5^ In addition, disparities in CI uptake exist among different patient cohorts: in the United Kingdom, for example, 74% of eligible children aged 0-3 years were estimated to have received implants compared to only 5% of eligible adults.^6^

Much of the current literature on cochlear implantation has focused on evaluating pre-defined health-related outcomes, such as communication/hearing scores and Quality of Life metrics.^7^ In contrast, there is a paucity of research examining patient experiences of cochlear implants, with survey- or interview-based studies often limited by small sample sizes of fewer than 100 participants.^8–12^ These studies may be poorly representative of the CI community, while their findings may be further constrained by the closed question-answer style format. Furthermore, existing qualitative research on CI user experience is often focused on identifying general positive benefits of CIs, such as improved user autonomy and psychological well-being,^8–12^ with limited analysis of patient concerns. In order to overcome these issues, it may be useful to explore non-traditional methods of evaluating the experiences and concerns of CI users, their families and other stakeholders.

Health researchers are increasingly utilising social media as a resource to generate data-driven insights.^13^ Social media offers multidirectional flow of communication between users, facilitating dynamic discussion on a variety of topics, including cochlear implantation. Despite the abundance and accessibility of health information on social media, only a handful of studies have sought to analyse the content of social media posts within the field of Otolaryngology. Among these is a qualitative study of 782 Twitter posts from parents of children who underwent a tonsillectomy and a thematic analysis of discussions around sudden sensorineural hearing loss in Reddit posts.^14,15^ Exploration of social media can give a unique insight into the experiences of patients that may not otherwise be shared with clinicians during traditional consultations. They also provide the opportunity to identify recurring worries or concerns from patients, reported ad-hoc in real time, and may be less prone to recall bias.

There are several active online CI communities on different social media platforms, where CI users and individuals interested in CIs can access and share information.^16^ Among these is the CI community that can be found on the online forum *Reddit*, a platform where people may anonymously converse by posting in community-based sub-forums known as subreddits. The CI subreddit, ‘r/Cochlearimplants’, currently has over 2400 members.^17^ Although Reddit is increasingly recognised as a valuable source of data in many other medical specialties,^18^ r/Cochlearimplants has not been examined thus far. An exploration of this subreddit, to identify common questions posed by prospective CI users, would allow clinicians to provide more detailed information to those wishing to consider implantation. Most importantly, understanding the experiences of existing and prospective CI users voiced in “non-clinician” controlled spaces may also give a different and perhaps more valid representation of their concerns, fears and what is valued compared to issues raised in clinician-user interactions.

In parallel to the emergence of social media as a resource for health research is the growing emphasis on using automated methods to analyse the potentially large volumes of data that are extracted. The field of natural language processing offers techniques to analyse text at scale, requiring considerably fewer resources and time compared to manual qualitative approaches.^19^ Topic modelling, where probabilistic algorithms are used to extract popular topics from a collection of documents, is one of these techniques and has been applied to a variety of bioinformatics tasks.^20^ However, the use of topic modelling techniques in Ear, Nose and Throat-related research is sparse, and there is limited manual evaluation of the accuracy of automatically-generated topics.^21,22^

The aims of the present study were therefore two-fold: Firstly, we sought to examine posts from the r/CochlearImplant Reddit forum to determine what were the most popular topics discussed by users. Secondly, we sought to investigate the utility of a state-of-the-art automated topic modelling technique applied to our dataset and assess its suitability for automated topic analysis in future Otolaryngology-related research.

## Methods

### Data collection

We used the Pushshift Multithread API Wrapper (PMAW)^23^ to access the Reddit API and extract all posts from the r/Cochlearimplants subreddit forum.^17^ Reddit was selected as the social media site for analysis due to its open accessibility and structured subreddit format. Ethical approval was not required as all data is in the public domain. Created on 1 March 2015, r/Cochlearimplants is a self-described ‘subreddit for cochlear implant users to share their stories, maintenance tips, advice, and have a general discussion about cochlear implants’,^17^ consisting of ∼2400 members. Data were retrieved on 11 November 2021. No date restrictions were imposed. The following details were extracted for each post: 1) original post content, 2) post comments, and 3) metadata for the original post and user. Original post and comment text were minimally preprocessed prior to topic modelling: the abbreviation ‘CI’ was expanded to ‘cochlear implant’ and ‘\n’ (denoting a new line) was removed.

### Topic modelling

We used a natural language processing approach to initially cluster Reddit posts (herein termed *documents*) into appropriate topic categories. Topics were identified using BERTopic, a topic modelling technique that utilises a Transformer model alongside a Class-based Term Frequency-Inverse Document Frequency (c-TF-IDF) procedure to cluster topics.^24^ Bidirectional Encoder Representations from Transformers (BERT) models represent state-of-the-art natural language processing models that use Transformer-based attention mechanisms to learn contextual relations between words.^25^ We used the sentence-BERT (all-MiniLM-L6-v2) model from the SentenceTransformers framework to compute sentence-level embeddings.^26^

In the BERTopic algorithm, first the Sentence-BERT model is used to generate document embeddings, then the dimensionality of embeddings is reduced by Uniform Manifold Approximation and Projection (UMAP).^27^ The reduced embeddings are then clustered by the HDBSCAN clustering algorithm into semantically similar groups of documents, and topics are extracted with c-TF-IDF. BERTopic is able to automate the number of topics generated, reducing topics sequentially until only topics which exceed a minimum similarity threshold of 0.915 remain. Any documents which do not cluster to a particular topic are placed in an ‘*uncategorised*’ group.

After applying the BERTopic algorithm to our dataset, the topics generated were inspected to confirm semantic similarity between documents. Overarching topic headings were then identified using a combination of the keywords extracted by BERTopic and manual identification of common themes. To verify the accurate classification of documents into each topic, two co-authors (CW and RL) manually and independently reviewed each document within a topic. Any documents deemed a poor fit with the given topic were identified and assigned to either a more appropriate topic or the *uncategorised* group. Disagreements between reviewers were resolved by consensus. The *uncategorised* group was subsequently inspected in duplicate and posts were manually assigned to either a new topic, an existing topic, or left as *uncategorised*. A summary of the initial clustering of documents into topics by BERTopic, and final topic categorisations, can be found in Supplementary Table 1.

### Statistical analysis

All analyses were performed on Python. To evaluate and compare the performance of BERTopic with human annotation, Cohen’s Kappa statistic was used to determine both a) inter-rater reliability between original BERTopic vs final human-modified Topic categorisation, and b) inter-human-rater reliability during manual categorisation of posts into Topics. Cohen’s kappa was calculated both on the entire (n = 987 posts) dataset and on a dataset consisting of the posts initially categorised by BERTopic (i.e excluding the 297 initially *uncategorised* posts).

## Results

We extracted 1061 Reddit posts from the r/CochlearImplants forum. Among these, 74 were found to have been censored or removed by Reddit and were therefore excluded, leaving 987 posts from 588 unique Reddit users for further analysis. The BERTopic algorithm categorised posts into 16 different Topics, with 297 posts initially left uncategorised. Following inspection of the *uncategorised* group, we manually created a further 6 Topics and either assigned each uncategorised post to a Topic or left it uncategorised (Table 1). An overview of the original (i.e BERTopic output) and final (human-modified) classification of Topics are shown in Figure 1.

**Table 1.** Overview of Topics from the r/CochlearImplant Reddit forum, ordered by prevalence. *topic manually curated; CI = cochlear implant

| Topic No. | Topic | Common themes | BERTopic most frequent words | Count (%) |
| --- | --- | --- | --- | --- |
| 1 | CI connectivity | Connectivity-related posts:<br>Connecting processor to phone; Android/Apple connectivity & functionality; connecting to smart watch; digital CI accessories | 'phone', 'bluetooth', 'clip', 'streaming', 'iphone', 'n', 'audio', 'connect', 'use', 'app' | 112 (11.3%) |
| 2 | Adults considering getting a CI | Questions from users considering getting a CI; majority of users aged >21 and generally in middle/older age generation | 'hearing', 'implant', 'cochlear implant', 'cochlear', 'ear', 'im', 'left', 'like', 'loss', 'people' | 107 (10.8%) |
| 3 | Surgery | Surgery-related posts:<br>3a) What to expect from the surgery; side effects after implantation; preparing for recovery (n = 84)<br>3b) Choosing where to go for surgery (n = 5) | 'surgery', 'tinnitus', 'pain', 'just', 'im', 'ear', 'implant', 'like', 'weeks', 'sleep' | 89 (9.0%) |
| 4 | Day-to-day living with a CI | Questions about day-to-day living with a CI:<br>What sports can you play? Are water sports (swimming, scuba diving) or horse riding or high impact sports okay? Can you use induction hobs, saunas? Can you go in an MRI scanner, near magnetic motors, airport scanners, near dusty environments? Difficulty wearing hats, hard hats, helmets, swimming goggles, glasses, VR headsets; keeping CI from falling off head | 'magnet', 'implant', 'head', 'cochlear', 'wear', 'cochlear implant', 'just', 'processor', 'im', 'use' | 85 (8.6%) |
| 5 | Sound quality | 5a) Activation stories/experiences and sound quality (n = 52)<br>5b) General posts on CI sound quality/performance (without mention of recent activation) (n = 25)<br>5c) Training hearing post-CI implantation (n = 5) | 'sounds', 'sound', 'im', 'hearing', 'hear', 'like', 'just', 'understand', 'activation', 'speech' | 82 (8.3%) |
| 6 | Supporting someone else | Supporting someone else considering/with a CI:<br>Often a family member supporting a young son/daughter; what to expect with a CI; should I look into a CI for son/daughter | 'deaf', 'cochlear', 'implant', 'hearing', 'cochlear implant', 'language', 'sign', 'just', 'like', 'daughter' | 73 (7.4%) |
| 7 | Gathering information | Gathering information about cochlear implants:<br>Including students asking the CI community for information for academic projects or general interest; survey requests; | 'cochlear', 'cochlear implants', 'cochlear implant', 'implant', 'hearing', 'implants', 'people', | 64 (6.5%) |
|  |  | fundraising requests; advertising other websites with CI information | 'information', 'hearing loss', 'like' |  |
| 8 | Choosing cochlear implant brand | Comparing attributes of different brands, tips on choosing | 'ab', 'cochlear', 'implant', 'processor', 'brand', 'im', 'like', 'cochlear implant', 'medel', 'advanced' | 52 (5.3%) |
| 9 | *Uncategorised* | Range of heterogeneous posts with no clear theme | 'implant', 'cochlear', 'hearing', 'cochlear implant', 'im', 'just', 'ear', 'like', 'sound', 'hear' | 47 (4.8%) |
| 10 | Sharing links to videos | Sharing links to CI-related videos | cochlear', 'cochlear implant', 'cochlear implants', 'implant', 'implants', 'video', 'page', 'news', 'cochlear implant surgery', 'implant surgery' | 45 (4.6%) |
| 11 | Battery and CI troubleshooting posts | 8a) Battery related: how to make batteries last longer, batteries draining too fast, charging issues, comparing battery life (n = 30)<br>8b) Issues with CI not turning on/not working properly (n = 11) | 'batteries', 'battery', 'rechargeable', 'charge', 'hours', 'power', 'cochlear', 'use', 'life', 'ones' | 41 (4.2%) |
| 12 | Insurance | Insurance-related posts:<br>How to pay for a CI; comparing insurance plans; insurance doesn't cover cost of CI | 'insurance', 'cover', 'cochlear', 'covered', 'cost', 'implant', 'cochlear implant', 'pay', 'im', 'plan' | 30 (3.0%) |
| 13 | Upgrading CI | Sharing/anticipating new releases; asking if upgrading is worth the cost; which version to get | 'nucleus', 'n', 'kanso', 'n', 'processor', 'upgrade', 'n', 'version', 'available', 'new' | 29 (2.9%) |
| 14 | Sharing other CI forums | Posts relating to other subreddit forums or other social media groups | 'deaf', 'discord', 'im', 'join', 'thank', 'cochlear', 'subreddit', 'app', 'cochlear implant', 'hi' | 26 (2.6%) |
| 15 | Music | Music-related: Listening to music with a CI | 'music', 'implant', 'sound', 'cochlear implant', 'cochlear', 'hearing', 'sounds', 'like', 'listen', | 24 (2.4%) |
|  |  |  | *'im'* |  |
| 16 | Coil issues | Coil/cable broken or not working | *'coil', 'cable', 'nucleus', 'upgrade', 'audiologist', 'freedom', 'im', 'processor', 'just', 'new'* | 16 (1.6%) |
| 17 | Miscellaneous symptoms in CI users | Collection of miscellaneous symptoms in long-time CI users, including tinnitus, hearing volume change with vaccination or upper respiratory tract infection and others | n/a* | 16 (1.6%) |
| 18 | External appearances/attitudes | 19a) Appearances/cosmetics of CI (n = 9)<br>19b) Attitudes from other people about CI; stigma experienced by CI users (n = 6) | n/a* | 15 (1.5%) |
| 19 | CI mappings | Discussing different programming settings/mappings | n/a* | 10 (1.0%) |
| 20 | CI failure | Failure of CIs and recalls | n/a* | 9 (0.9%) |
| 21 | Joining the CI community | General posts sharing CI journey so far and desire/excitement to join the CI online community | n/a* | 8 (0.8%) |
| 22 | CI-related movies | Discussing CI or hearing-loss related movies:<br>In particular, the movie 'Sound of Metal' | n/a* | 4 (0.4%) |
| 23 | Internal cochlear implants | Discussing CIs with no external components; entire implant is internal | *'internal', 'cochlear', 'implants', 'cochlear implants', 'implant', 'cochlear implant', 'hearing', 'think', 'know', 'wouldnt'* | 3 (0.3%) |

**Figure 1.**
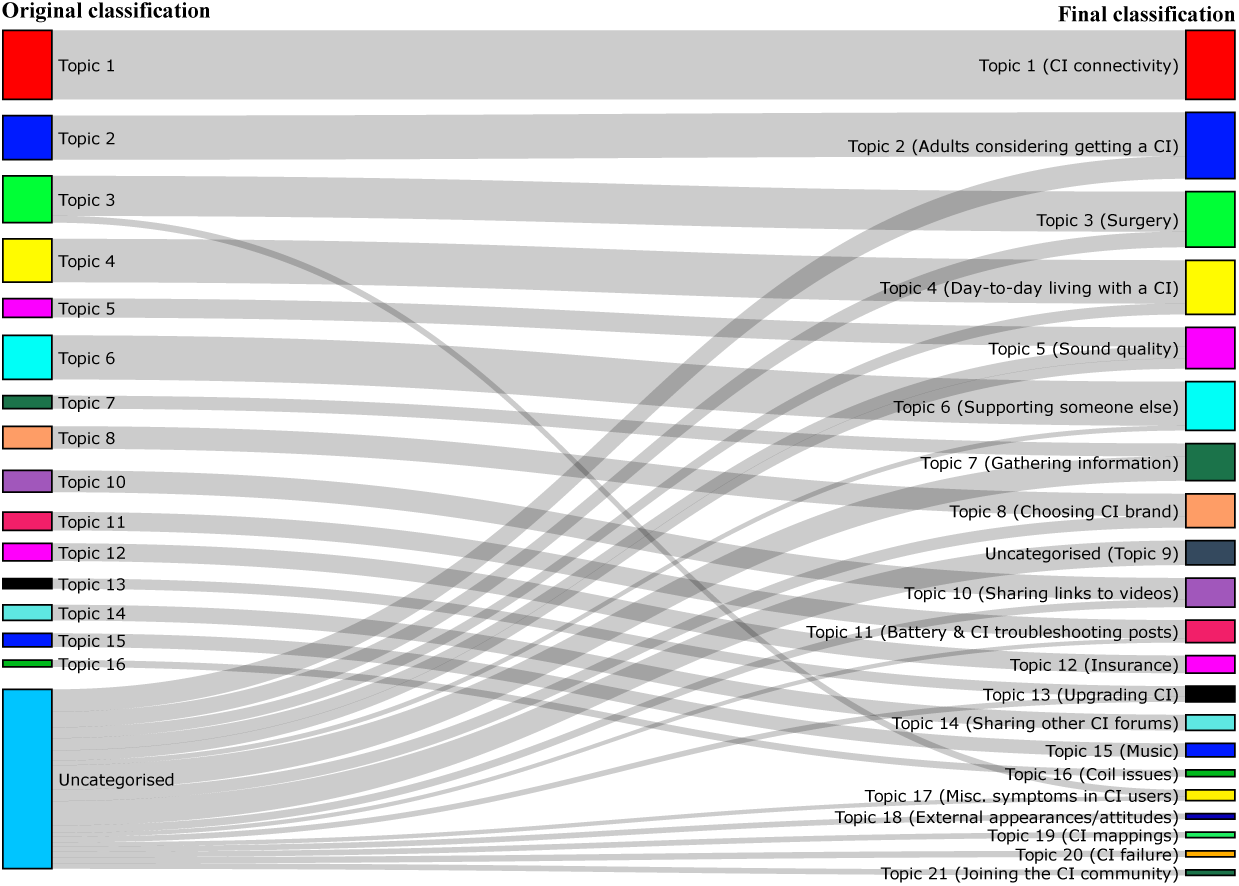
Overview of the original (i.e BERTopic output) and final (human-modified) classification of Topics. N.B: Topic changes of ≤5 in quantity have been excluded for ease of viewing. A full breakdown of Original vs Final Topics can be found in S1 Table.

### Topic model performance

Cohen’s kappa for inter-rater reliability between the original (machine-labelled) and final (human-checked) Topic classification was 0.62. This compares to a Cohen’s kappa of 0.72 for inter-rater reliability between the two human annotators. When the initially *uncategorised* group was excluded from the analysis, Cohen’s Kappa improved to 0.85 (original vs final Topic classification) and 0.84 (inter-human classification).

### Common themes

The evolution of topic popularity among Reddit users over time is shown in Figure 2. The most popular topics related to CI connectivity (Topic 1, n = 112), adults considering getting a CI (Topic 2, n = 107), surgery-related posts (Topic 3, n = 89) and day-to-day living with a CI (Topic 4, n = 85) (Table 1). CI connectivity posts (Topic 1) commonly mentioned questions about connecting to other devices or related to digital CI accessories such as smart watches, baby monitors and phone clips. Surgery-related posts (Topic 3) typically sought advice on what to expect from the surgery, common side effects after implantation, and how best to prepare for recovery post-op. Among the posts discussing day-to-day living with a CI, the issues raised varied greatly and included questions on which sports CI users can play and whether certain environments or appliances are safe, alongside complaints about difficulty wearing headgear with a CI (e.g helmets, hats, masks) and from users wishing to prevent their CI from falling off.

**Figure 2.**
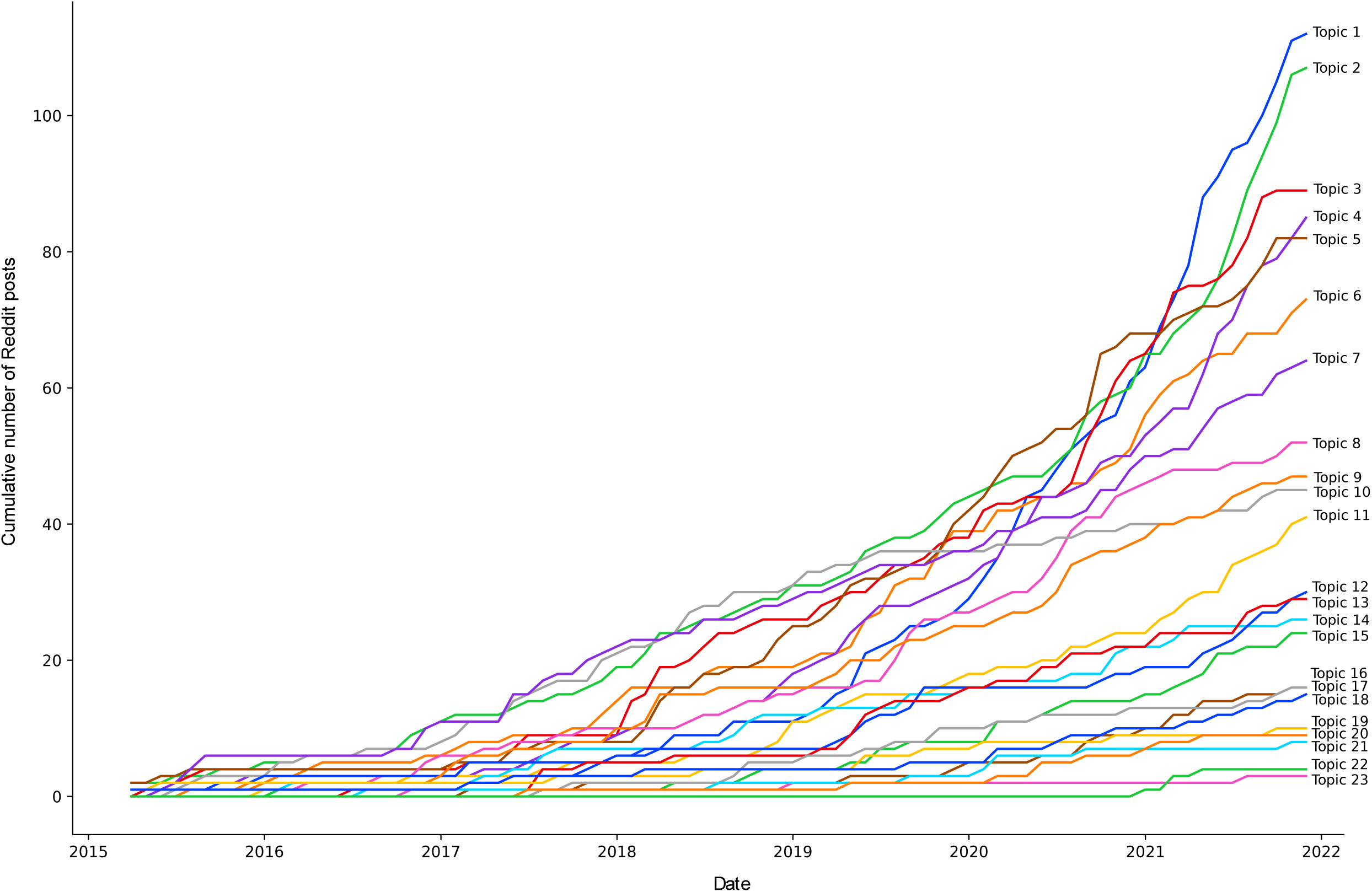
Evolution of topic popularity among r/CochlearImplants posts over time

Two distinct groups of posts emerged among users broadly discussing whether to get a CI. A larger group (Topic 2, n = 107) consisted of adults considering getting a CI and looking for more information about it. While not all posts in this group mentioned the user’s age, those which did were predominantly middle to older age individuals who were considering getting a CI later in life. This contrasts a second group (Topic 6, n = 73) of posts which related to supporting someone else and considering them for a CI. Here, often a family member was seeking advice when supporting a young son/daughter who is being considered for a CI. A third group also consisted of posts gathering information about CIs (Topic 7, n = 64), but these requests were for general interest or academic projects rather than people wishing to explore getting a CI.

A total of 82 posts (Topic 5) covered discussions of CI sound quality, while a further 24 related to listening to music (Topic 15). The BERTopic algorithm was successfully able to distinguish between posts discussing the differences between CI brands (Topic 8, n = 52) from posts evaluating whether to upgrade to newer versions of the same brand (Topic 13, n = 29). Several Topics were identified that covered different aspects of the overarching theme of *CI malfunction*; these include: a) Battery and CI troubleshooting posts (Topic 11, n = 41), where posters discussed battery draining/charging issues and CIs not working properly; b) Coil issues (Topic 16, n = 16) and c) CI recalls/failures (Topic 20, n = 9). Meanwhile, 30 posts discussed issues of paying for CIs and insurance-related complaints or questions (Topic 12).

Lastly, the remaining topics covered a broad range of CI-related content. These include sharing links to videos (n = 45), sharing other CI forums (n = 26), discussing miscellaneous symptoms (n = 16), external appearances and attitudes towards CIs (n = 15), CI mappings (n = 10) and CI-related movies (n = 4).

### Topic engagement

There were a total of 8102 comments associated with the 987 posts in our dataset; each post attracted a median of 6 comments (interquartile range: 3 – 12). Topics with the most frequent number of comments included Topic 8 (Choosing cochlear implant brand; 12.9 comments on average), Topic 2 (Adults considering getting a CI; 12.2 comments on average), and Topics 6 (Supporting someone else), 22 (CI-related movies), 20 (CI failure) and 18 (Joining the CI community), with an average of 11 comments each (Table 2). Conversely, the top 5 topics with the most upvoted posts were Topic 21 (Joining the CI community; average score = 4.8), Topic 10 (Sharing links to videos; average score = 3.0), Topic 18 (External appearances/attitudes; average score = 2.9), Topic 14 (Sharing other CI forums; average score = 2.8) and Topic 5 (Sound quality; average score = 2.7) (Table 3).

**Table 2.** Topic engagement by mean number of comments

| Topic No. | Topic | Mean No. of comments |
| --- | --- | --- |
| 8 | Choosing cochlear implant brand | 12.9 |
| 2 | Adults considering getting a CI | 12.2 |
| 6 | Supporting someone else | 11.6 |
| 22 | CI-related movies | 11.5 |
| 20 | CI failure | 11.3 |
| 21 | Joining the CI community | 10.8 |
| 18 | External appearances/attitudes | 9.9 |
| 3 | Surgery | 9.8 |
| 5 | Sound quality | 9.7 |
| 15 | Music | 9.2 |
| 4 | Day-to-day living with a CI | 8.7 |
| 17 | Miscellaneous symptoms in CI users | 8.1 |
| 11 | Battery and CI troubleshooting posts | 8.0 |
| 23 | Internal cochlear implants | 7.7 |
| 16 | Coil issues | 7.6 |
| 13 | Upgrading CI | 7.2 |
| 1 | CI connectivity | 6.4 |
| 12 | Insurance | 6.1 |
| 19 | CI mappings | 6.0 |
| 7 | Gathering information | 4.3 |
| 9 | Uncategorised | 2.6 |
| 14 | Sharing other CI forums | 2.0 |
| 10 | Sharing links to videos | 1.2 |

**Table 3.**
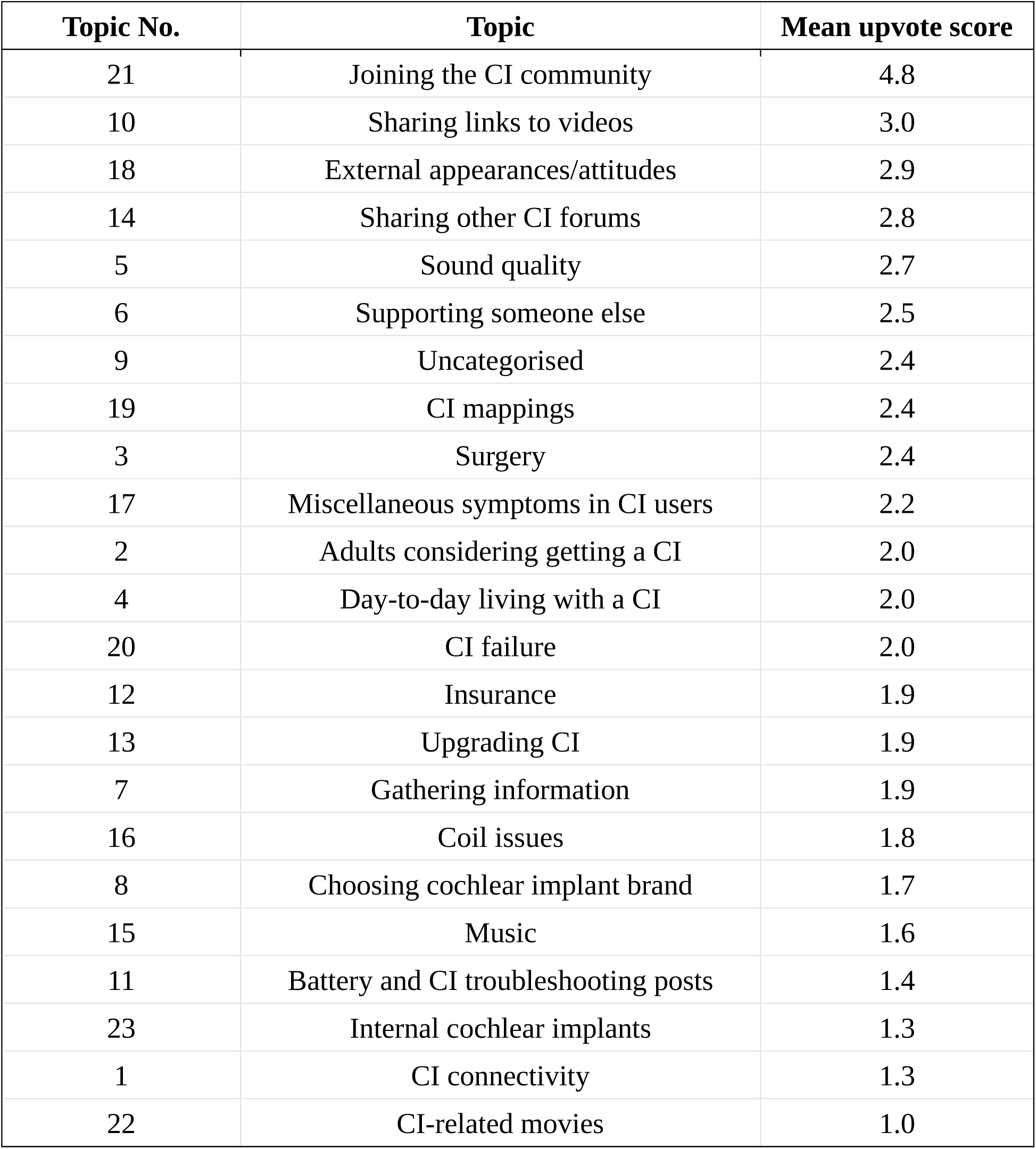
Topic engagement by mean upvote score

## Discussion

We have identified a variety of common themes from discussions posted by members of the Reddit cochlear implant community. These range from CI connectivity and day-to-day living with a CI to queries about choosing a CI brand and post-op expectations. Although the extant literature evaluating CI user experiences is limited, previous studies have suggested that the cost of devices, fear of complications from surgery, and worries about sound quality/listening to music are among the barriers to CI uptake.^28–31^ Similar topics were identified in our analysis, with posts relating to surgery and sound quality consistently ranking among the top five most popular topics.

However, the prevalence of other topics in our analysis differed from the results of prior studies. For instance, in one survey of parental reported shortcomings of CIs, a frequent need for repairs, especially to cables, was reported among 30% of respondents.^32^ Meanwhile, separate studies have found almost 20% of CI users reported battery problems as a concern, with short battery life and the risk of batteries suddenly going dead a recurring problem.^8,33^ While these issues were also identified in our analysis, they were much less prevalent, representing only 1.6% (cable/coil issues) and 4.2% (battery issues) of posts. It is possible that the reported frequency of these topics has been skewed by the small number of participants in previous studies when compared to the present one, reflecting the potential value of using non-traditional sources such as social media to obtain larger sample sizes for data collection.

Our analysis identified several frequently mentioned themes which have not been previously reported. Among these was the topic of CI connectivity, which contained the highest number of posts in our dataset. As technology has evolved, new digital accessories that wirelessly connect CIs to everyday devices have been developed, seeking to improve sound quality and user experience across different settings.^34,35^ Our results show that this greater connectivity is of increasing interest among the CI community, especially over the last two years. Many users posting about this topic reported issues with either connecting between different devices (Apple vs Android etc) or with the initial setup of their device. This suggests that standardising CI accessory compatibility across a range of different devices is a key area for future improvement. Meanwhile, examining posts about day-to-day living with a CI revealed some of the common questions raised by prospective CI users concerned about how implantation may affect their lives. Queries as to the feasibility of different sports were frequently posted, alongside reports on the difficulty of keeping CIs from falling off while wearing various forms of headwear. As newer CI models are developed, it is important that these issues are considered by CI manufacturers in order to improve device usability.

When exploring the level of engagement of individual posts, discussions regarding which cochlear implant brand to go for received the highest number of comments. This reflects both the wide variety of available device options and uncertainty as to which device might perform better over others,^36^ and suggests that greater support for patients when choosing a CI device may be required. In addition, posts from adults either considering getting a CI, or supporting someone else with a CI, received high numbers of comments. These findings demonstrate the value of the r/CochlearImplant forum as a medium through which users might engage with each other to receive advice and support. Analysis of the most upvoted topics further confirms this, with posts that discussed external appearances/attitudes associated with CIs, or reported users’ CI journey so far, consistently receiving higher ‘upvotes’ compared to other topics.

This study also represents the first reported validation of automated topic modelling methods to categorise social media posts within the field of Ear, Nose and Throat surgery. We sought to manually verify the appropriateness of automated topic categorisation and found similar Cohen’s kappa scores for inter-rate reliability between human-machine and human-human categorisation of posts, ranging from 0.62 to 0.85. This suggests that, although some posts are inherently more difficult than others to categorise into a particular topic, automated topic modelling is able to perform this categorisation with similar accuracy to human annotators. Based on these findings, we propose that using natural language processing is a viable approach to analyse large datasets of unstructured text relating to other areas of Otolaryngology.

### Limitations

Our study has several limitations. It is difficult to establish how representative Reddit is in relation to the online CI community or the cochlear implant community as a whole. It is possible that some prospective/actual CI users and family either do not have access to Reddit and other social media forums, or choose not to engage with other CI users on online platforms. Although the exact proportion of CI users belonging to the Reddit community is uncertain, a previous systematic review has confirmed that a strong presence of CI users and individuals interested in CIs exists online from a variety of social media sources.^16^ Due to the pseudonymous nature of Reddit, we also could not obtain detailed demographic information about the age, gender or ethnicities of users posting about a particular topic. Future research must therefore consider differences among minority groups to ensure that patient experiences and concerns are equitably identified across the population.

## Conclusion

To our knowledge, this is the first study to explore the content of social media discussions amongst the online cochlear implant community. Analysis of social media posts allowed us to identify common attitudes, experiences and concerns among patients either living with, or seeking, a cochlear implant. Importantly, these perspectives may not otherwise have been expressed during a patient’s interactions with their otolaryngologist, or captured by researchers using existing survey-based techniques. Our validation of natural language processing methods to categorise topics in this setting further shows that automated analysis of similar Otolaryngology-related content is both a viable and accurate alternative to manual qualitative approaches.

## Data Availability

All data produced in the present study are either contained in the manuscript or available upon reasonable request to the authors

