## Supplementary Table 1 for "Exploring patient experiences and concerns in the online Cochlear Implant community: a natural language processing approach"

| **Original Topic Categorisation** | **Final Topic Categorisation** | **Count** |
| --- | --- | --- |
| 1 | 1 | 102 |
| 1 | 13 | 3 |
| 1 | 12 | 2 |
| 1 | 15 | 1 |
| 2 | 2 | 65 |
| 2 | 9 | 4 |
| 2 | 18 | 2 |
| 2 | 6 | 1 |
| 2 | 13 | 1 |
| 2 | 4 | 1 |
| 2 | 5 | 1 |
| 3 | 3 | 59 |
| 3 | 17 | 10 |
| 3 | 5 | 2 |
| 3 | 7 | 1 |
| 3 | 2 | 1 |
| 4 | 4 | 64 |
| 4 | 16 | 4 |
| 4 | 2 | 1 |
| 4 | 3 | 1 |
| 5 | 5 | 28 |
| 5 | 5 | 5 |
| 5 | 2 | 3 |
| 5 | 5 | 2 |
| 5 | 19 | 1 |
| 5 | 3 | 1 |
| 5 | 14 | 1 |
| 6 | 6 | 65 |
| 6 | 7 | 2 |
| 6 | 2 | 2 |
| 6 | 5 | 2 |
| 6 | 1 | 1 |
| 6 | 12 | 1 |
| 6 | 4 | 1 |
| 6 | 5 | 1 |
| 7 | 7 | 19 |
| 7 | 2 | 1 |
| 8 | 8 | 33 |
| 8 | 13 | 1 |
| 8 | 2 | 1 |
| 8 | 10 | 1 |
| 9 | 7 | 36 |
| 9 | 9 | 36 |
| 9 | 2 | 33 |
| 9 | 3 | 23 |
| 9 | 5 | 18 |
| 9 | 8 | 17 |
| 9 | 4 | 16 |
| 9 | 5 | 15 |
| 9 | 10 | 11 |
| 9 | 20 | 9 |
| 9 | 13 | 8 |
| 9 | 19 | 8 |
| 9 | 21 | 8 |
| 9 | 18 | 8 |
| 9 | 6 | 7 |
| 9 | 17 | 6 |
| 9 | 11 | 6 |
| 9 | 1 | 5 |
| 9 | 3 | 5 |
| 9 | 18 | 4 |
| 9 | 22 | 4 |
| 9 | 15 | 3 |
| 9 | 5 | 3 |
| 9 | 11 | 3 |
| 9 | 16 | 2 |
| 9 | 14 | 2 |
| 9 | 12 | 1 |
| 10 | 10 | 32 |
| 10 | 9 | 1 |
| 11 | 11 | 27 |
| 11 | 11 | 3 |
| 11 | 1 | 1 |
| 11 | 4 | 1 |
| 12 | 12 | 26 |
| 12 | 4 | 1 |
| 13 | 13 | 15 |
| 13 | 1 | 2 |
| 13 | 8 | 2 |
| 13 | 5 | 1 |
| 14 | 14 | 23 |
| 14 | 9 | 4 |
| 14 | 7 | 3 |
| 14 | 10 | 1 |
| 15 | 15 | 20 |
| 15 | 5 | 3 |
| 15 | 5 | 1 |
| 16 | 16 | 10 |
| 16 | 11 | 2 |
| 16 | 13 | 1 |
| 16 | 18 | 1 |
| 23 | 7 | 3 |
| 23 | 23 | 3 |
| 23 | 9 | 2 |
| 23 | 1 | 1 |
| 23 | 19 | 1 |
| 23 | 4 | 1 |
|  | **Total** | **987** |

**Table S1**. Comparison of original (BERTopic output) vs final (human-modified) topic categorisation, ordered by *Original Topic Categorisation* then *Count*
